# Targetable *ERBB2* mutation status is an independent marker of adverse prognosis in estrogen receptor positive, *ERBB2* non-amplified primary lobular breast carcinoma: a retrospective *in silico* analysis of public datasets

**DOI:** 10.1101/2020.01.24.20018622

**Authors:** Sasagu Kurozumi, Mansour Alsaleem, Cíntia J. Monteiro, Kartikeya Bhardwaj, Stacey E. P. Joosten, Takaaki Fujii, Ken Shirabe, Andrew R. Green, Ian O. Ellis, Emad A. Rakha, Nigel P. Mongan, David M. Heery, Wilbert Zwart, Steffi Oesterreich, Simon J. Johnston

**Affiliations:** Nottingham Breast Cancer Research Centre, School of Medicine, University of Nottingham, UK; Department of General Surgical Science, Gunma University Graduate School of Medicine, Gunma, Japan; Gene Regulation and RNA Biology Laboratory, School of Pharmacy, University of Nottingham, UK; School of Veterinary Medicine and Science, University of Nottingham, UK; Division of Oncogenomics, Oncode Institute, Netherlands Cancer Institute, Amsterdam, Netherlands; Womens Cancer Research Center, UPMC Hillman Cancer Center and Magee-Women Research Institute, Pittsburgh, PA, USA

**Keywords:** Lobular, breast cancer, *ERBB2*, HER2, mutation, prognosis, therapeutic biomarker, adjuvant

## Abstract

**Background:** Invasive lobular carcinoma (ILC) accounts for 10-15% of primary breast cancers and is typically estrogen receptor alpha positive (ER+) and *ERBB2* non-amplified. Somatic mutations in *ERBB2/3* are emerging as a tractable mechanism underlying enhanced human epidermal growth factor 2 (HER2) activity. We tested the hypothesis that therapeutically targetable *ERBB2/3* mutations in primary ILC of the breast associate with poor survival outcome in large public datasets.

**Methods:** We performed *in silico* comparison of *ERBB2* non-amplified cases of ER+ stage I-III primary ILC (*N*=279) and invasive ductal carcinoma (IDC, *N*=1,301) using METABRIC, TCGA and MSK-IMPACT information. Activating mutations amenable to HER2-directed therapy with neratinib were identified using existing functional data from *in vitro* cell line and xenograft experiments. Multivariate analysis of 10-year overall survival (OS) with tumor size, grade and lymph node status was performed using a Cox regression model. Differential gene expression analyses by *ERBB2* mutation and amplification status was performed using weighted average differences and an *in-silico* model of response to neratinib derived from breast cancer cell lines.

**Results:** ILC tumors comprised 17.7% of all cases in the dataset but accounted for 47.1% of *ERBB2*-mutated cases. Mutations in *ERBB2* were enriched in ILC *versus* IDC cases (5.7%, *N*=16 *vs*. 1.4%, *N*=18, *p*<0.0001) and clustered in the tyrosine kinase domain of HER2. *ERBB3* mutations were not enriched in ILC (1.1%, *N*=3 *vs*. 1.8%, *N*=23; *p*=0.604). Median OS for patients with *ERBB2*-mutant ILC tumors was 66 months *versus* 211 months for *ERBB2* wild-type (*p*=0.0001), and 159 *vs*. 166 months (*p*=0.733) for IDC tumors. Targetable *ERBB2* mutational status was an independent prognostic marker of 10-year OS – but only in ILC (hazard ratio, HR=3.7, 95% CI 1.2–11.0; *p*=0.021). Findings were validated using a novel *ERBB2* mutation gene enrichment score (HR for 10-year OS in ILC=2.3, 95% CI 1.04–5.05; *p*=0.040).

**Conclusions:** Targetable *ERBB2* mutations are enriched in primary ILC and their detection represents an actionable strategy with the potential to improve patient outcomes. Biomarker-led clinical trials of adjuvant HER-targeted therapy are warranted for patients with *ERBB2*-mutated primary ILC.

## Background

Invasive lobular carcinoma of the breast (ILC) accounts for 10-15% of all breast cancer with an estimated 250,000 cases per year worldwide [1-4]. Nearly all cases of ILC derive from luminal cells that express estrogen receptor alpha but lack E-Cadherin (*CDH1*) expression or *ERBB2* amplification [5].

Clinical evidence suggests that despite favorable prognostic indicators, e.g. ER+ and/or progesterone receptor positive, low Ki67 proliferation index and HER2-status, patients with ILC have similar or worse long-term outcomes compared to those with invasive ductal carcinoma (IDC, otherwise known as invasive carcinoma of no special type) [5, 6]. Adjuvant treatment of ER+ ILC with letrozole, an aromatase inhibitor, may be superior to tamoxifen, and ILC cells demonstrate resistance to tamoxifen *in vitro* [7, 8]. However, patients with ILC are treated according to identical protocols as those with IDC [9, 10].

HER2-targeted therapy is indicated for patients whose tumors are HER2+ by immunohistochemistry (IHC) or, if IHC is equivocal, where *ERBB2* is amplified as detected by *in situ* hybridization (ISH) [11, 12]. Recent evidence indicates that in *ERBB2* non-amplified breast cancer, somatic mutation of *ERBB2* (*ERBB2*mut) and/or *ERBB3* (*ERBB3*mut) may provide an alternative mechanism for upregulation of HER2 activity that is therapeutically tractable using second generation HER2 tyrosine kinase inhibitors such as neratinib [13, 14].

The epidermal growth factor (EGF) family of receptor tyrosine kinases (HER1-4) are activated by ligand-dependent homo-/heterodimerisation and regulate cellular proliferation and tumor progression [15, 16]. In *ERBB2* amplified cells, the oncogenic effect of HER2 is mediated by heterodimerisation with HER3 in a ligand-independent manner [17]. Thus HER3 is necessary for HER2 oncogenic activity, and both HER2 and HER3 are therapeutic targets in *ERBB2* amplified breast cancer [18]. Mutations in *ERBB2* cluster in the tyrosine kinase and extra-cellular domains of HER2 and exert their oncogenic effects by activating tyrosine kinase activity or increasing HER2 dimerisation, respectively [13]. *In vitro* studies of HER2 activity in cell line and xenograft models identified 13 mutations (listed in the Methods) that enhanced proliferation and/or demonstrated growth inhibition with the irreversible HER2/EGFR tyrosine kinase inhibitor, neratinib [13]. Mutations in HER3, a critical binding partner for HER2, have been shown to promote ligand (EGF)-independent transformation of breast epithelial cells only in the presence of kinase-active HER2 [19]. This indicates that known oncogenic mutations in *ERBB3*, e.g. G284R and E928G, may also be therapeutically targetable *via* HER2 inhibition [19].

*ERBB2* mutations have previously been linked with worse prognosis in *CDH1*-altered ILC: a study of ILC cases in the TCGA dataset (*N*=169) found that *ERBB2* mutations (*N*=6) occurred exclusively in *CDH1-*altered tumors (*N*=100) [20]. However, prognostic data on the 6 *ERBB2*mut cases were limited to 2 patient events for both disease-free survival and OS analyses. The overall rate of *ERBB2*mut in this study of primary ILC was 3.6% (*N*=6 out of 169). A study of relapsed *CDH1*-mutated ILC found *ERBB2*mut in 18% of cases (*N*=4 of 22), suggesting further acquisition of *ERBB2* mutations in *CDH1*-altered ILC due to the selective pressure of treatment [21].

To demonstrate the potential clinical benefit of targeting low frequency somatic mutations, prognostic analyses using large clinical datasets are required. In the MA12 trial comprising 328 premenopausal patients with ER+ primary breast cancer of all histological subtypes, non-silent *ERBB2* mutations occured in 5.2% of patients (*N*=17) and were adversely prognostic of OS (*p*=0.0114) [22]. A study of 5,605 cases of relapsed breast cancer found *ERBB2*mut in 2.4% of cases (*N*=138), of which 20% (*N*=27 of 138) were in ILC tumors [23]. However, neither of these larger studies stratified clinical outcome by histological subtype.

An ILC-specific study of 630 cases of primary ILC found *ERBB2* and *ERBB3* mutations in 5.1% and 3.6% of tumors respectively [24]. Comparison of cases of ER+, HER2-ILC from the same study (*N*=371) with cases of ER+, HER2-IDC from TCGA (*N*=338) indicated significant enrichment of both *ERBB2* and *ERBB3* mutations in ILC (4.3% and 3.5% in ILC *vs*. 1.5% and 0.6% in IDC) [24]. The study reported limited statistical evidence of a time-dependent effect of *ERBB2* mutational status associated with short-term breast cancer-specific survival. However, confirmation in datasets including patients with long-term follow-up and decoupling of activating from silent mutations is needed.

We hypothesized that low-frequency somatic mutations in *ERBB2* and *ERBB3* are enriched in ER+, *ERBB2* non-amplified primary ILC cases and may have demonstrable prognostic effect. We tested these hypotheses by mining a combined dataset of the three largest primary breast cancer series with data on tumor *ERBB2* and *ERBB3* mutational status, gene expression, clinicopathological features and patient survival outcomes. Our overall goal was to determine the association between targetable *ERBB2/3* mutations and survival in ILC, and thereby provide evidence for a clinically actionable strategy to improve outcomes for patients with ILC.

### Patients and methods

#### Patients and outcome measures

Genomic and clinical outcome data associated with tumor samples from patients with primary breast cancer in TCGA 2015 (*N*=817), METABRIC 2012 and 2016 (*N*=2,509) and MSK-IMPACT 2018 (*N*=918) were accessed online *via* CBioportal [25-27]. From these datasets, ER+ and HER2-cases of stage I-III ILC and IDC with both clinical outcome and mutational data called from next-generation sequencing (NGS) analyses were selected (*N*=1,580). Cases of mixed or non-ILC/IDC histology, ER-negative/undetermined, HER2+/undetermined, carcinoma-*in-situ*, and stage 4/undetermined were excluded. For TCGA and MSK datasets, HER2 status was determined by IHC (positive/negative) or, where IHC was indeterminate, by ISH assessment of *ERBB2* amplification, in line with standard clinical practice [11]. For METABRIC cases, HER2 status was determined using the Affymetrix SNP6 copy number inference pipeline.

The primary outcome measure, available in all datasets, was OS. Variables included *ERBB2* and *ERBB3* mutational status. *ERBB2*mut status was subcategorized as oncogenic or uncharacterized by cross-reference with existing data that identified mutations targetable by HER2-inhibition: G309A/E, S310F, L755S, del755-759, S760A, D769H, D769Y, V777L, P780ins, V842I, R896C [13]. Cases were denominated onc*ERBB2*mut if tumors harbored at least one oncogenic *ERBB2* mutation.

Clinical and NGS mutation data were integrated with clinicopathological features including histological subtype, lymph node (LN) status and tumor size and grade. Normalized gene expression data were publicly available for METABRIC (Illumina HT12 microarray) and TCGA (RNA-seq) datasets. Baseline clinicopathological characteristics are summarized in Table 1.

**Table 1:** Baseline clinicopathological characteristics of the combined cohort. *Cases of ILC/IDC histology, stage I-III, ER+ and HER2-status with clinical outcome and mutational data were selected *via* CBioportal. * *at last follow-up.

|  |  | METABRIC<br>(N=702) |  | TCGA<br>(N=330) |  | MSK-IMPACT<br>(N=548) |  | TOTAL<br>(N=1580) |  |
| --- | --- | --- | --- | --- | --- | --- | --- | --- | --- |
|  |  | N | % | N | % | N | % | N | % |
| **Histology\*** | ILC | 76 | 10.8 | 100 | 30.3 | 103 | 18.8 | 279 | 17.7 |
|  | IDC | 626 | 89.2 | 230 | 69.7 | 445 | 81.2 | 1301 | 82.3 |
| **Age** | <50 yrs | 125 | 17.8 | 87 | 26.4 | 190 | 34.7 | 402 | 25.4 |
|  | ≥50 yrs | 577 | 82.2 | 243 | 73.6 | 358 | 65.3 | 1178 | 74.6 |
| **Menopause** | Pre- | 125 | 17.8 | 90 | 27.2 | 234 | 42.7 | 449 | 28.5 |
|  | Post- | 577 | 82.2 | 219 | 66.4 | 309 | 56.4 | 1105 | 69.9 |
|  | unknown | 0 | 0 | 21 | 6.4 | 5 | 0.9 | 26 | 1.6 |
| **Stage\*** | I | 238 | 33.9 | 67 | 49.3 | 270 | 49.3 | 575 | 36.4 |
|  | II | 418 | 59.5 | 188 | 34.3 | 188 | 34.3 | 794 | 50.3 |
|  | III | 46 | 6.6 | 75 | 16.4 | 90 | 16.4 | 211 | 13.4 |
| **Tumor size** | <20mm | 215 | 30.6 | 96 | 29.1 | 330 | 60.2 | 641 | 40.6 |
|  | ≥20mm | 487 | 69.4 | 234 | 70.9 | 218 | 39.8 | 939 | 59.4 |
| **Tumor grade** | 1 | 61 | 8.7 | 49 | 14.8 | 51 | 9.3 | 161 | 10.2 |
|  | 2 | 344 | 49.0 | 190 | 57.6 | 191 | 34.9 | 725 | 45.9 |
|  | 3 | 268 | 38.2 | 81 | 24.5 | 288 | 52.6 | 637 | 40.3 |
|  | unknown | 29 | 4.1 | 10 | 3.0 | 18 | 3.3 | 57 | 3.6 |
| **Follow-up** | <5 yrs | 137 | 19.5 | 261 | 82.8 | 454 | 82.8 | 852 | 53.9 |
|  | 5-10 yrs | 195 | 27.8 | 61 | 11.7 | 64 | 11.7 | 320 | 20.3 |
|  | ≥10 yrs | 370 | 52.7 | 8 | 5.5 | 30 | 5.5 | 408 | 25.8 |
| **Status** | Alive** | 310 | 44.2 | 299 | 90.1 | 494 | 90.1 | 1103 | 69.8 |
|  | Deceased | 392 | 55.8 | 31 | 9.9 | 54 | 9.9 | 477 | 20.3 |

### Statistical analysis

For analysis of binary somatic mutation status, combined cohort analysis was performed. Enrichment for cases with mutations in candidate genes (*ERBB2/3*mut) by histological subtype (ILC *vs*. IDC) was determined by χ^2^ test for association between categorical variables. For ILC and IDC separately, Kaplan-Meier (KM) survival curves stratified by mutation status were compared using Logrank and generalized Wilcoxon tests. Multivariate analysis of OS was performed using a Cox-regression model. Covariates included tumor grade, size (<20mm or ≥20mm) and LN status (positive or negative). Tumor grade was classified as low (grade 1-2) or high (grade 3).

To derive a novel gene expression signature of HER2 activity that accounted for the effect of potentially targetable *ERBB2*mut in *ERBB2* non-amplified tumors, we applied a weighted average difference (WAD) method to gene expression data in cases from the METABRIC 2012 (*N*=1,980) and TCGA 2015 (*N*=817) cohorts [25, 28, 29]. Gene expression in *ERBB2*mut cases (*N*=38, selected by *ERBB2* non-amplified status and patient age>50) was compared with that in a similar number of *ERBB2* wild-type cases (*N*=79, selected by *ERBB2* non-amplified status, grade>1, stage>I, patient age>50). This was repeated for onc*ERBB2*mut cases (*N*=23) using the same comparator and selection criteria. To incorporate the effect of HER2 activity *via ERBB2* amplification, the overlap of differentially expressed genes DEGs shared by both comparisons (*ERBB2*mut and onc*ERBB2*mut *vs. ERBB2* wild-type) with DEGs from a further comparison of *ERBB2* amplified (*N*=247) *versus* non-amplified (*N*=1,733) cases in METABRIC was calculated. Finally, to incorporate the downstream phenotype (HER2 status), the overlap of this list with DEGs from a comparison of clinical HER2+ *versus* HER2-cases in TCGA was calculated.

Multiple gene expression signatures of HER2 activity have been derived using cell line models and patient tumors [30-33]. We compared our novel gene signature with the HER2 activity signature established by Desmedt *et al* [31] with respect to its ability to detect potentially targetable *ERBB2*mut cases in our ILC/IDC dataset. This was achieved by multivariate regression modeling of response to neratinib for each gene signature using breast cancer cell line pharmacogenomic data from the BROAD Institute, accessed online *via* the CellMinerCDB portal [34, 35]. The Pearson coefficient for each significantly correlated signature gene was used to calculate normalized signature scores for each METABRIC case in the current dataset. To validate the prognostic effect of *ERBB2* in ILC, cases were then stratified by gene signature score (upper *vs*. lower quartiles) and the signatures compared by histological subtype using a Cox regression model of 10-year OS.

## Results

### Clinicopathological landscape of the combined cohort

To detect prognostic effects of low frequency mutations and add to the existing body of literature on ILC-specific mutational drivers, a combined cohort of three public datasets was collated. We first evaluated the implications of combining cases of ILC and IDC from potentially disparate datasets.

In the combined cohort, long-term follow-up of patients is dominated by the largest dataset (METABRIC, N=702: mean follow-up 133 months) whilst early events are enriched by the two smaller datasets (TCGA and MSK, N=878: mean follow-up 33 months). Compared to the smaller datasets with shorter follow-up (TCGA and MSK), patients in METABRIC were more likely to be over 50 years of age and have T1 tumours (<20mm diameter), but no significant difference in grade or LN status was found (see Table 2). As the principle skew in the combined dataset is towards longer follow-up in METABRIC cases, OS analysis was limited to 10 years – thus providing a clinically meaningful endpoint for all patients, irrespective of age.

**Table 2:** Comparison of long *vs*. short follow-up cohorts: METABRIC (largest dataset, long follow-up) *vs*. TCGA and MSK (combined smaller datasets, short follow-up). Significant difference was found with respect to age and tumor size, but not tumor grade or LN status. Table excludes “unknown” cases for each variable.

| | | METABRIC<br>(N=702) | | TCGA AND MSK<br>(N=878) | | $\chi^2$-test |
| --- | --- | --- | --- | --- | --- | --- |
|  |  | N | % | N | % |  |
| **Age** | <50 years | 125 | 17.8 | 277 | 31.5 | $p<0.00001$ |
|  | ≥50 years | 577 | 82.2 | 601 | 68.5 |  |
| **Tumor size** | <20mm | 215 | 30.6 | 426 | 48.5 | $p<0.00001$ |
|  | ≥20 mm | 487 | 69.4 | 452 | 51.5 |  |
| **Tumor grade** | 1-2 | 405 | 60.2 | 481 | 56.6 | $p=0.158$ |
|  | 3 | 268 | 39.8 | 369 | 43.4 |  |
| **LN status** | Negative | 370 | 53.9 | 489 | 55.8 | $p=0.457$ |
|  | Positive | 316 | 46.1 | 387 | 44.2 |  |

### *ERBB2* mutations are enriched in primary ILC and cluster in the HER2 tyrosine kinase domain

We next assessed the prevalence of *ERBB2/3*mut in our dataset of ILC and IDC (*N*=1,580). Overall prevalence of *ERBB2*mut was 2.2% (*N*=34). *ERBB2*mut was enriched in ILC, with prevalence of 5.7% (*N*=16) *vs*. 1.4% in IDC (*N*=18) (*p*<0.0001). In contrast, prevalence of *ERBB3*mut was lower (1.6% overall, *N*=26), and there was no enrichment of *ERBB3*mut in ILC *vs*. IDC (1.1%, *N*=3 *vs*. 1.8%, *N*=23; *p*=0.604). Due to the small number of cases in ILC, further analysis of *ERBB3*mut was not performed.

In ILC, *ERBB2*mut clustered in the tyrosine kinase domain of HER2 (15 out of 16 cases; 93.8%). Of these, the majority have been characterized as oncogenic and potentially targetable using neratinib (onc*ERBB2*mut, 11 out of 15 kinase domain mutations in ILC; 73.3%). In IDC, all kinase domain mutations were known oncogenic (*N*=8) and non-characterized mutations were distributed evenly across protein domains (see Figure 1). The most frequently occurring *ERBB3*mut coded for E928G, which lies in the tyrosine kinase domain of HER3 (*N*=7 out of 26; 27%).

**Figure 1:**
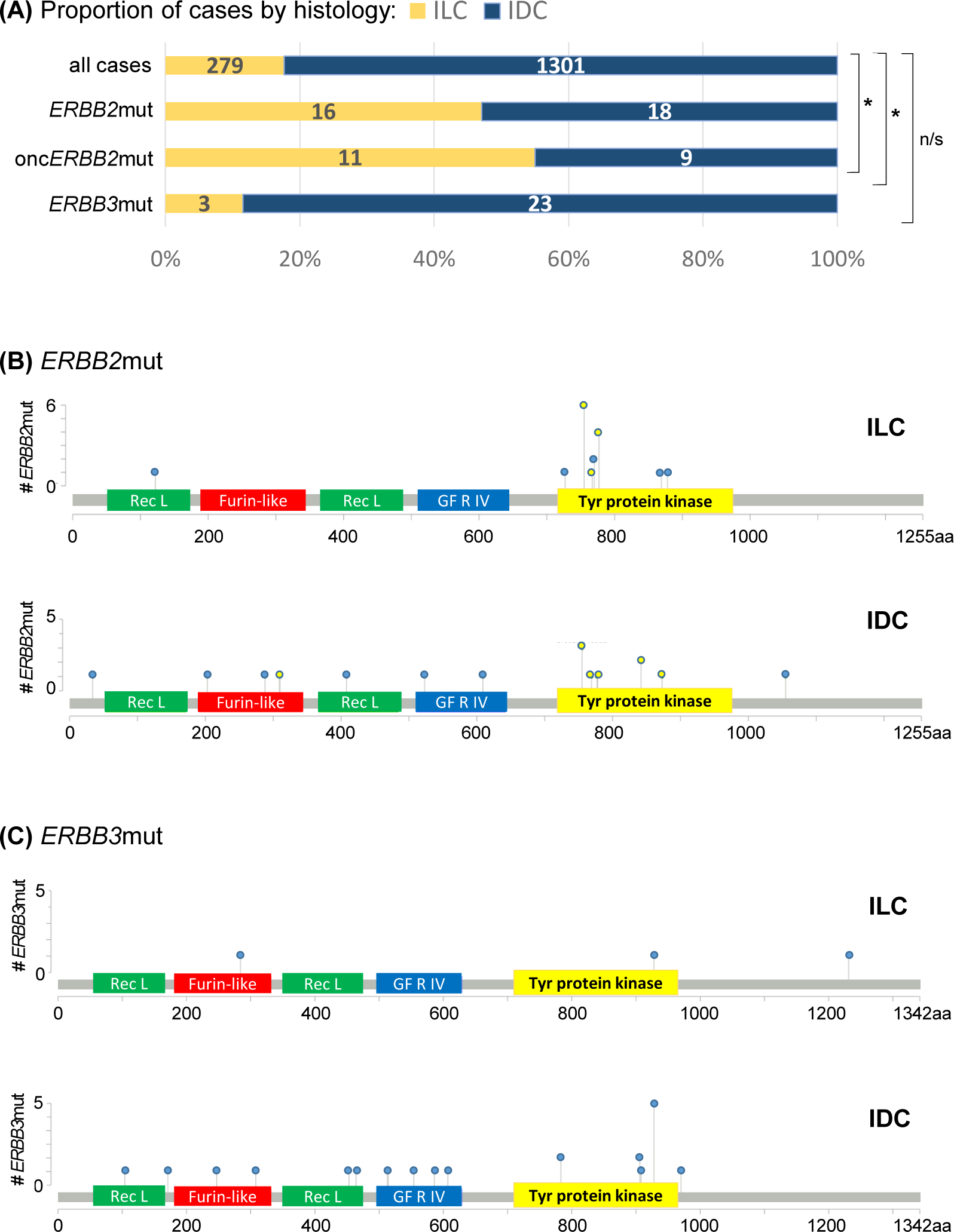
Rate of *ERBB2* and *ERBB3* mutations and their spatial distribution on HER2/3 in ILC and IDC. *ERBB2*mut were found to be **(A)** enriched in ILC (yellow bar) *vs*. IDC (blue bar) and **(B)** clustered in the tyrosine kinase domain of HER2; **(C)** *ERBB3*mut occurred at lower frequency, with the high-frequency outlier in IDC coding for known oncogenic HER3 kinase domain alteration E928G (*N*=6). Y-axes show the number of cases harboring at least one *ERBB2/3* mutation at a specific amino acid (aa) of HER2/3, shown along the x-axes. Yellow-filled circles indicate onc*ERBB2*mut. Extracellular domains of HER2/3: Receptor L, Furin-like and Growth Factor Receptor IV; intracellular: tyrosine protein kinase. \**p*<0.001; n/s = not significant.

### *ERBB2* mutation is an adverse prognostic indicator of survival for patients with primary ILC

Using our meta-cohort of 1,580 cases, we next assessed *ERBB2*mut as a prognostic marker of OS in ILC (*N*=279) and IDC (*N*=1,301). Median duration of patient follow-up was 50 months (range 0 – 351 months). In patients with ILC, median OS was significantly shorter if tumors were *ERBB2*mut positive (inclusive of oncogenic and uncharacterized mutations) *versus ERBB2*mut negative (66 *vs*. 211 months, *p*=0.0001). In contrast, there was no significant difference in OS for *ERBB2*mut cases of IDC (159 *vs*. 166 months, *p*=0.733) (see Figure 2).

**Figure 2:**
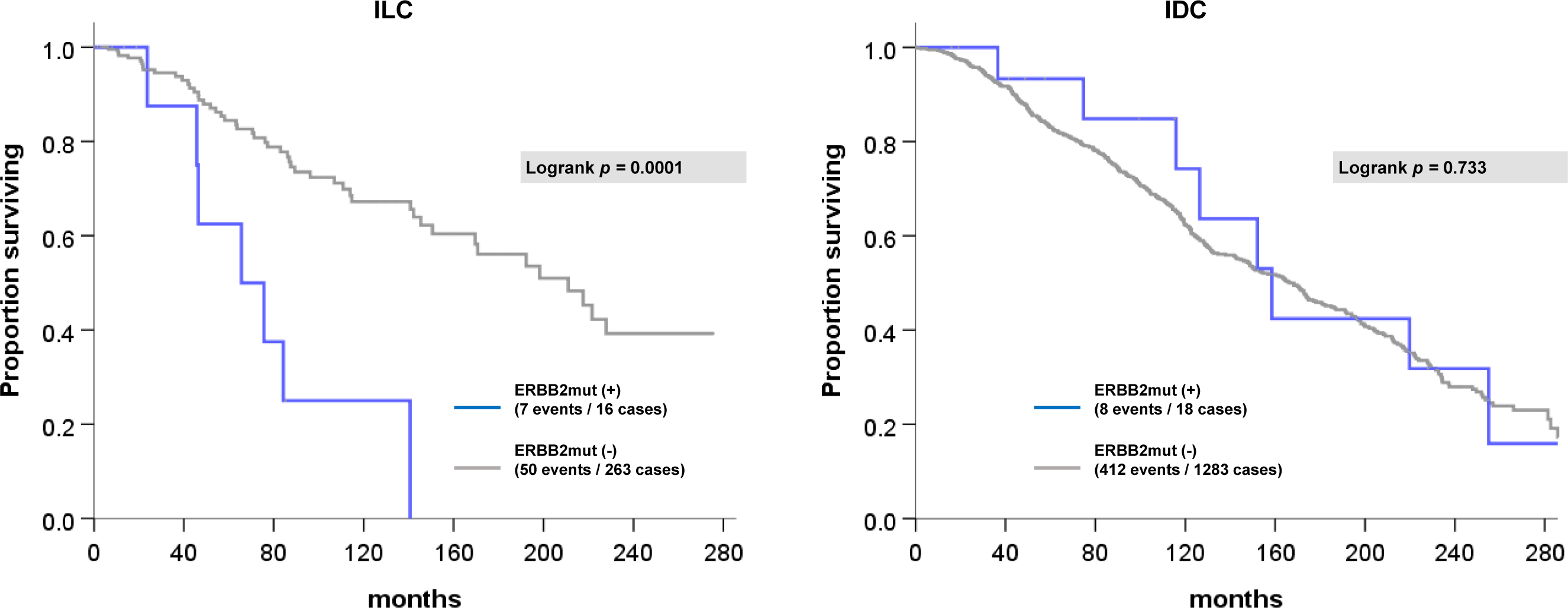
OS by *ERBB2* mutational status in ILC (left) and IDC (right). Gray line indicates *ERBB2* wild-type cases; blue line indicates cases with at least one *ERBB2*mut.

*ERBB3*mut status was not a significant prognostic indicator of OS in cases of IDC (HR=1.46, 95% CI 0.65 – 3.28; *p*=0.359). There were no events (patient deaths) at ten years of follow-up in the three cases of *ERBB3*mut ILC.

### Targetable *ERBB2* mutation status is an independent adverse prognostic marker of 10-year overall survival in ER+, *ERBB2* non-amplified ILC

To test the effect of therapeutically actionable *ERBB2* mutations on a clinically relevant endpoint, we stratified 10-year OS by onc*ERBB2*mut status. In ER+, *ERBB2* non-amplified ILC, onc*ERBB2*mut status was prognostic of 10-year OS independently of LN status, tumor grade and size (HR 3.65, 95% CI 1.21 – 11.00; *p*=0.021, see Figure 3).

**Figure 3.**
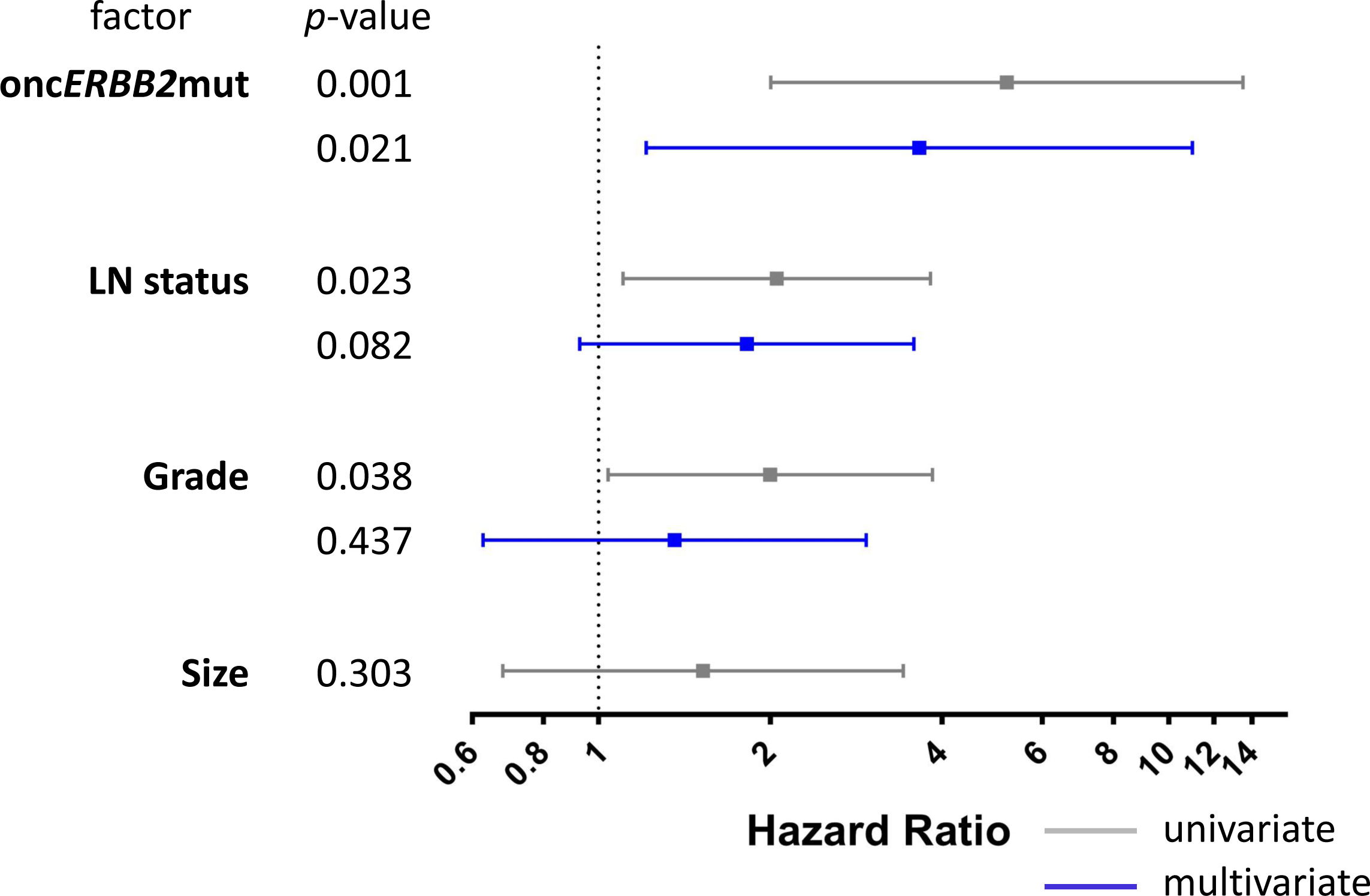
Univariate and multivariate analyses of 10-year OS in *N*=279 cases of ER+, *ERBB2* non-amplified ILC. Gray square dot indicates hazard ratio (HR) in univariate analysis and the gray bar indicates the 95% confidence interval (CI). Significant prognostic variables in univariate analyses (where 95% CI does not span HR=1) are included in multivariate analysis, shown in blue. For each variable, cases with unknown values are excluded from the analysis.

Unselected *ERBB2*mut status (including oncogenic and uncharacterized mutations) was also adversely prognostic of 10-year OS in univariate analysis (HR 3.66, 95% CI 1.54 – 8.73; *p*=0.003), but in contrast to characterized onc*ERBB2*mut this prognostic effect was not independent of LN stage, tumor grade and / or size.

### A novel gene signature of targetable HER2 activity using *ERBB2*mut status

Since existing gene signatures of HER activity derive from HER2 status (by IHC and/or ISH) we generated a novel gene signature incorporating DEGs in *ERBB2*mut and onc*ERBB2*mut cases, as described in the Methods and outlined in Figure 4. A list of up and down-regulated *ERBB2* “mutant” DEGs (*N*=20) was generated by combining the overlap between DEGs for METABRIC *ERBB2* amplified *versus* non-amplified, *ERBB2* mutated *versus* wild-type (*ERBB2*mut and onc*ERBB2*mut separately) and TCGA HER2+ *versus* HER2-(Figure 4A, and see Additional file 1 for supplemental tables S1-3 with all gene lists).

**Figure 4:**
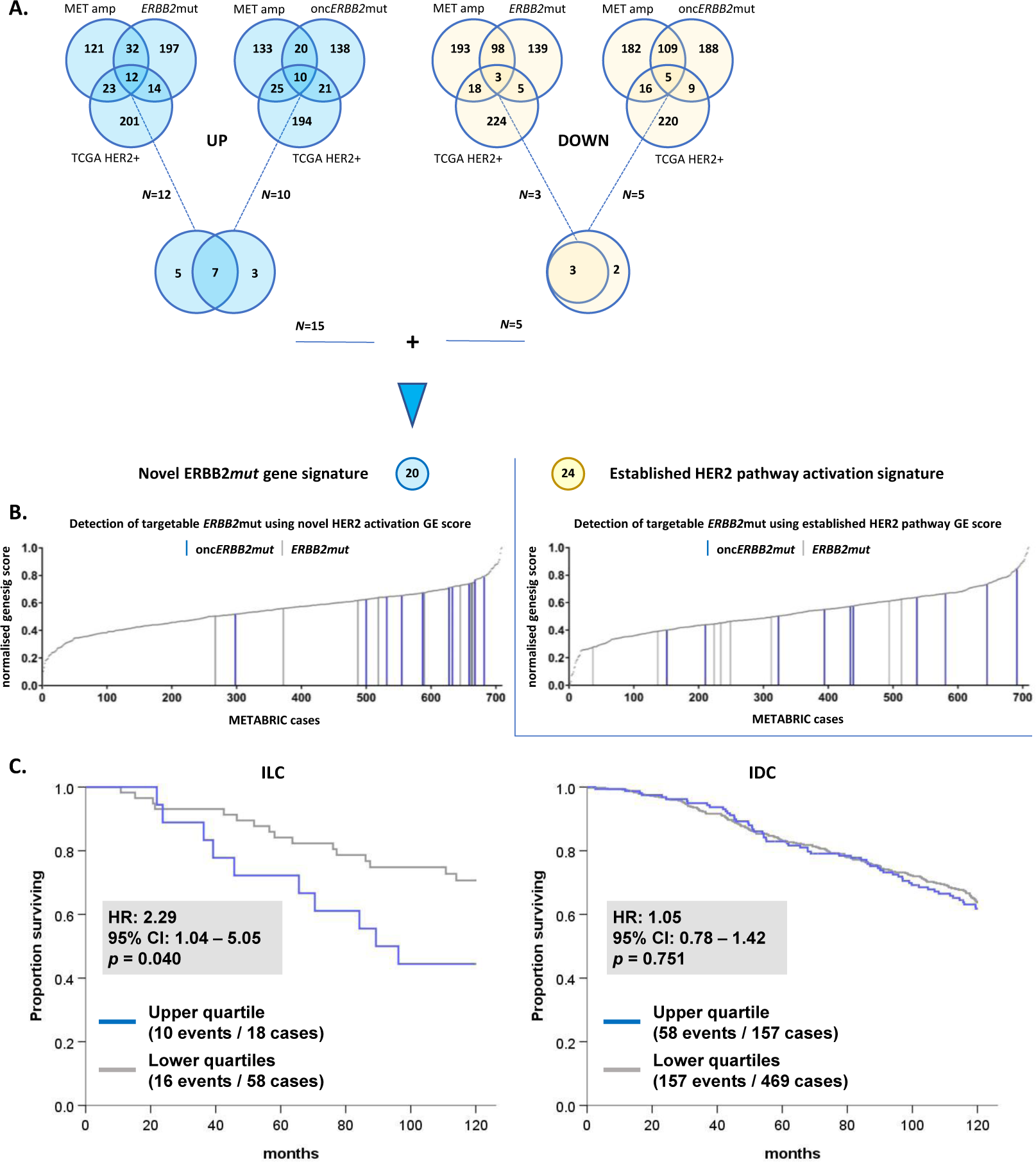
A novel gene signature of HER2 activity incorporating *ERBB2*mut, *ERBB2* amplification and clinical HER2 status. **(A)** Generation of a 20-gene signature of HER2 activity. The upper Venn diagrams show the overlap between the top 500 DEGs by WAD score for METABRIC amplified *vs*. non-amplified, *ERBB2* mutant *vs*. wild-type, and TCGA HER2+ *vs*. HER2-. Up-regulated DEGs are shaded blue and down-regulated DEGs yellow. *ERBB2*mut and onc*ERBB2*mut *vs*. wild-type are analysed separately, and the overlap combined in the lower Venn diagrams. **(B)** Comparison with an established 24-gene signature of HER pathway activation [31], using a gene signature (genesig) score derived from multivariate analysis of response to neratinib in breast cancer cell lines. Cases with *ERBB2*mut (gray lines) and onc*ERBB2*mut (blue lines) clustered in the upper quartile of normalized genesig scores for the novel signature but not the established signature. **(C)** 10-year OS analysis of cases in the current study stratified by histological subtype and novel genesig score (upper *vs*. lower quartiles) indicates that *ERBB2*mut-associated DEGs are prognostic in ILC but not IDC. GE = geneset enrichment.

As shown in Figure 4B, *ERBB2*mut and onc*ERBB2*mut clustered in the upper quartile of the novel gene signature score in cases of ER+, HER2-ILC/IDC from METABRIC. In contrast, *ERBB2*mut and onc*ERBB2*mut cases did not cluster when scored by the established gene signature of HER2 pathway activation but were evenly distributed across the cohort. Both the novel and established HER2 pathway gene signatures were predictive of response to neratinib in multivariate analysis using a pharmacogenomic breast cancer cell line model (Pearson R=0.9, *p*<1e^-14^ for both; see Additional file 2 for the original plots from the CellMinerCDB online portal). The regression model used incorporates drug response and gene expression data from *N*=36 breast cancer cell lines [35, 36]. Our findings using this model verify that the novel signature derived from DEGs in *ERBB2*mut *versus* wild-type cases reflects oncogenic HER2 activity due to *ERBB2*mut. This suggests that a gene signature may have value in predicting the presence of *ERBB2*mut in HER2-breast cancer. Finally, Figure 4C demonstrates that the novel gene signature score (stratified into upper *vs*. lower quartiles) was adversely associated with 10-year OS (HR=2.3, 95% CI 1.04–5.05; *p*=0.040), thus providing mRNA level validation of *ERBB2*mut as an adverse prognostic marker.

## Discussion

In this study, we mined clinical and NGS data from the largest clinical cohorts with data in the public domain to test whether there was an ILC-specific association of *ERBB2* mutations with OS. In our meta-cohort of ER+, *ERBB2* non-amplified cases of ILC and IDC, we found that *ERBB2* mutations are enriched ILC, cluster in the functional kinase domain of HER2, and robustly associate with adverse clinical outcomes – independently of known prognostic clinicopathological features including LN status and tumor grade. In contrast, there was no ILC-specific enrichment of *ERBB3*mut.

Compared to the largest previous study with data on *ERBB2* mutational status in primary ILC (*N*=371) [24], the current study found higher frequency of *ERBB2*mut (5.7% *vs*. 4.3%) but lower frequency of *ERBB3*mut (1.1% *vs*. 3.5%). Hazard proportionality in the study by Desmedt *et al* 2016 suggested that *ERBB2* mutational status had a time-dependent effect associated with short-term risk of breast cancer relapse [24]. The current study adds data on the prognostic effect of specific *ERBB2* mutations that were previously shown in cell line, *in vitro* and xenograft studies to respond to existing clinical compounds such as trastuzumab or neratinib [13]. We demonstrated that the status of these therapeutically targetable *ERBB2* mutations is an independent adverse prognostic marker of overall survival in a large cohort of patients with ILC.

Findings from the current study imply that targeted sequencing of ILC tumors for *ERBB2* mutations may be an actionable and viable strategy to improve outcomes for patients with primary ILC by providing a biomarker for HER-targeted therapy in the adjuvant setting. Based on worldwide ILC incidence of 250,000 [1-4], 10% rate of ER- and/or HER2-status in ILC, and 5% rate of potentially targetable *ERBB2*mut in ER+, *ERBB2* non-amplified ILC, we conservatively estimate that 11,250 additional ILC patients per year who have *ERBB2*-mutated tumors may benefit from HER2-targeting therapy.

Existing clinical studies of HER2-targeting therapy are underway in advanced breast cancer. For example, neratinib (alone or in combination with fulvestrant) was tested in phase II study for patients with advanced, *ERBB2* non-amplified, *ERBB2*mut breast cancer (clinical trial registration number NCT01670877). Of the 16 patients who had evaluable tumor responses, *N*=2 had partial response by RECIST criteria (13%). Out of 14 patients with tumors harboring onc*ERBB2*mut, *N*=5 (36%) derived clinical benefit, defined as stable disease or partial/complete response by RECIST criteria [37]. Given that the patients in this trial had received multiple prior lines of treatment, it is conceivable that patients with *ERBB2* non-amplified primary tumors harboring onc*ERBB2*mut will also derive benefit from (neo)adjuvant HER2-targeted therapy such as neratinib.

In the current cohort of primary ER+, HER2-ILC and IDC, *ERBB2* mutations in ILC tumors clustered almost exclusively in the active HER2 tyrosine kinase domain (*N*=15 of 16). Our retrospective clinical outcome data indicates an ILC-specific adverse survival association for patients with *ERBB2* non-amplified tumors harboring potentially targetable *ERBB2* mutations. This primary finding was validated by existing mRNA data *via* a novel gene signature of HER2 activity, which linked *ERBB2*mut with response to neratinib. Subject to independent external validation, a gene signature of HER2 activity could have clinical application as a biomarker of response to HER2-targeted therapy and may be more cost-effective than targeted NGS in the detection of *ERBB2*mut in primary ILC.

Taken together, these new findings suggest that focusing early phase studies of HER2-inhibition (e.g. with neratinib) in patients with *ERBB2* non-amplified, *ERBB2*mut primary ILC may be an effective strategy to demonstrate feasibility and clinical benefit in the (neo)adjuvant setting.

Limitations of the current study include cohort size, quality of clinical outcome data and bias in long-term follow-up towards cases from the largest dataset (METABRIC). Statistical power is limited by the small number of patient deaths in cases of ILC and IDC with *ERBB2*mut (*N*=7 and 8 respectively). However, the clear deviation of the KM plot of OS in ILC stratified by *ERBB2*mut (Figure 2) is reflected in a low type I error of one in 10,000 (*p*=0.0001). In contrast, there was no ILC-specific enrichment of *ERBB3*mut in our database. However, in IDC there was enrichment of the known oncogenic HER3 kinase domain mutation, E928G. To demonstrate the prognostic effect of mutations affecting HER3, larger datasets will be required.

The effect of incomplete clinical outcome data on the primary endpoint (OS) is difficult to quantify but is accounted for statistically in Cox regression modelling by censoring patients at last follow-up. Patients in METABRIC presented at older age, which is associated with more indolent, less biologically aggressive breast cancer [38]. This implies an underestimation of the adverse impact of *ERBB2* mutations on 10-year survival outcomes in the current study. Since the METABRIC dataset did not have complete HER2 IHC or reverse-phase protein array data, it was not possible to associate activating *ERBB2*mut directly with downstream HER2 protein expression. Instead, we generated a gene signature incorporating clinical HER2 status to validate our findings.

## Conclusions

Targetable kinase domain *ERBB2* mutations are enriched in primary ILC and their detection by targeted sequencing or validated gene signature surrogate may provide an actionable strategy to improve patient outcomes. Biomarker-led clinical trials of adjuvant HER2-targeted therapy to treat breast tumors with activating *ERBB2* mutations are warranted in HER2-primary ILC.

## Data Availability

The datasets generated and/or analysed during the current study are available via the CBioportal online portal (http://www.cbioportal.org) and in the Genome-Phenome Archives EGAS00000000098 and phs000178.

https://ega-archive.org/studies/EGAS00000000098

https://ega-archive.org/studies/phs000178

## List of abbreviations

CI: Confidence interval
DEG: Differentially expressed gene
EGF: Epidermal growth factor
ER: Estrogen receptor alpha
*ERBB2/3*mut: *ERBB2/3* mutant
HER2/3: Human epidermal growth factor receptor 2 / 3
HR: Hazard ratio
IDC: Invasive ductal carcinoma of the breast
IHC: Immunohistochemistry
ILC: Invasive lobular carcinoma of the breast
ISH: *In situ* hybridization
KM: Kaplein-Meier
LN: Lymph node
NGS: Next generation sequencing
onc*ERBB2*mut: Oncogenic *ERBB2* mutant
OS: Overall survival
RECIST: Response evaluation criteria in solid tumors
WAD: Weighted average difference

## Declarations

### Ethics approval and consent to participate

Ethics approvals and consent for each cohort (METABRIC, TCGA and MSK-IMPACT) was obtained in the original studies.

### Consent for publication

Not applicable

### Availability of data and materials

The datasets generated and/or analysed during the current study are available *via* the CBioportal online portal (http://www.cbioportal.org) and in the Genome-Phenome Archives EGAS00000000098 and phs000178 (https://ega-archive.org/studies/EGAS00000000098 and https://ega-archive.org/studies/phs000178).

### Authors’ contributions

SK, DMH, WZ, SO and SJJ conceived, designed and supervised the study. SK, MA, CM, KB, NPM and SJJ analysed and interpreted the data. SK, WZ, SO and SJJ prepared the manuscript. ARG, IOE and EAR had supervisory and administrative roles. SJ, TF and KS had consultative roles in study design. All authors were involved in manuscript preparation and reviewing for submission, gave their final approval and agreed to be accountable for all aspects of the work.

## Acknowledgements

Not applicable

## Additional files

**Additional file. 1** Supplemental tables: **(S1)** list of genes in the novel (*N*=20) and established (Desmedt *et al* [31], *N*=24) gene signatures of HER2 activity; **(S2 and S3)** top 500 DEGs by WAD score stratified into genes that are up-regulated (S2) and down-regulated (S3) for METABRIC *ERBB2* amplified *vs*. non-amplified (*ERBB2*amp), METABRIC *ERBB2*mut *vs*. wild-type (*ERBB2*mut) and onc*ERBB2*mut *vs*. wild-type (onc*ERBB2*mut), and TCGA HER2+ *vs*. HER2-(TCGA HER2+).

**Additional file 2**. Supplemental figure showing the correlation between the observed response of breast cancer cell lines (*N*=36) to neratinib and the response predicted by expression of (A) novel (*N*=20) and (B) established (Desmedt *et al* [31], *N*=24) gene panels. Plots were generated using CellMinerCDB online portal and cell line data from the BROAD Institute [34, 35].

